# Bioenergetic dysregulation in the basal ganglia and cerebellum of patients with premanifest and manifest Huntington’s disease

**DOI:** 10.64898/2026.01.12.26343913

**Authors:** Jannik Prasuhn, Maximilian G. Ködderitzsch Mertins, Marta M. Pokotylo, Joke-Lina Aßmann, Leon van Well, Julia Henkel, Jan Uter, Sebastian Loens, Alexander Münchau, Norbert Brüggemann

## Abstract

**Background:** Huntington’s disease (HD) is an autosomal dominant neurodegenerative disorder characterized by progressive motor, cognitive, and psychiatric symptoms. While early striatal degeneration is a well-established hallmark, emerging evidence points to broader network-level dysfunction involving the cerebellum and profound alterations in mitochondrial energy metabolism. However, *in vivo* studies systematically examining region-specific bioenergetic changes across disease stages are scarce.

**Methods:** Using ^31^Phosphorus magnetic resonance spectroscopic imaging (^31^P-MRSI), we quantified metabolite ratios and absolute concentrations of alpha adenosine triphosphate (ATP-α), phosphocreatine (PCr), and inorganic phosphate (Pi) in the anterior basal ganglia and cerebellum of 31 patients with HD (15 manifest, 16 premanifest) and 19 mutation-free healthy controls (HC).

**Results:** Basal ganglia (ATP-α + PCr)/Pi and ATP-α/Pi were significantly elevated in premanifest (+10.0% and +13.9%) and manifest patients (+16.4% and +19.2%) compared to HC. Contrarily, HEP levels demonstrated a stage-dependent pattern, with ATP-α + PCr and ATP-α elevated in premanifest (+7.7% and +3.9%) and reduced in manifesting patients (−7.6% and −12.8%). All metabolite ratios showed no correlation with clinical scores but were inversely associated with subcortical atrophy, particularly in the caudate (r = −0.352, p = 0.013), putamen (r = −0.391, p = 0.005), and globus pallidus (r = −0.335, p = 0.012).

**Conclusions:** These findings reveal stage-dependent, region-specific alterations in HEP metabolism in patients with HD. The observed changes in the brain energy metabolism precede motor symptoms and are linked to structural atrophy rather than symptomatic burden, supporting the potential of ^31^P-MRSI as a sensitive in vivo biomarker of bioenergetic dysfunction in HD.

## INTRODUCTION

Huntington’s disease (HD) is a progressive neurodegenerative disorder caused by an expanded CAG repeat mutation in the huntingtin (*HTT)* gene on chromosome 4.(1) The resulting mutant huntingtin (mHTT) protein disrupts numerous cellular processes, including transcription, axonal transport, and mitochondrial dynamics.(2) Mutation carriers inevitably develop the disease, progressing from a clinically silent premanifest phase to a stage of progressive overt motor, cognitive, and psychiatric symptoms.(3,4) A hallmark of HD is early and selective striatum degeneration, which disrupts the integrity of cortico-striatal-thalamic loops and underlies the core motor and psychiatric features of the disease.(5,6)

Beyond the striatum, recent studies implicate additional brain regions, particularly the cerebellum, in HD pathophysiology.(7) Traditionally viewed as a motor coordination center, the cerebellum is increasingly recognized for its involvement in cognitive and affective processes and its dynamic interactions with basal ganglia circuits.(8) In movement disorders such as Parkinson’s disease, dystonia, and HD, cerebellar engagement may reflect compensatory activity aimed at modulating impaired striato-cortical output; however, its potential adaptive versus pathogenic role in HD remains unclear.(8) Whether this engagement represents adaptive plasticity or contributes to network-level dysfunction in HD remains an open question.

Mitochondrial dysfunction has emerged as a central mechanism in HD, affecting energy production, redox balance, calcium handling, and apoptotic signaling. Neurons in the striatum and cerebellum are particularly vulnerable to impaired bioenergetics due to their high energy demands and complex synaptic architecture.(9–11) *In vivo* ^31^phosphorus magnetic resonance spectroscopic imaging (^31^P-MRSI) detects adenosine triphosphate (ATP), phosphocreatine (PCr), and inorganic phosphate (Pi), among other high-energy phosphates (HEP), which serve as key indicators of mitochondrial function and cellular energy. ATP levels reflect the immediately available energy for neuronal processes, whereas PCr reflects the reserve levels of energy substrates that are mobilized in response to fluctuating metabolic demands. Pi is produced as a result of ATP hydrolysis, with its elevated levels suggesting elevated energy consumption or impaired oxidative phosphorylation. Furthermore, ratios such as (ATP + PCr)/Pi, ATP/Pi, and PCr/Pi served as markers of cellular phosphorylation potential and energetic efficiency. While alterations in mitochondrial respiration, ATP synthesis, and PCr buffering capacity have been demonstrated in preclinical models and postmortem tissue, *in vivo* evidence from human studies remains limited.(9,12–16) To address this gap, we employed *in vivo* ^31^P-MRSI to investigate ATP, PCr, and Pi levels in the subcortical gray matter and cerebellum of individuals with premanifest and manifesting HD. By comparing metabolic profiles across disease stages and with mutation-free healthy controls (HC), we aimed to determine whether HD is associated with region-specific changes in energy metabolism and whether such changes emerge before clinical manifestation.

## METHODS

### Recruitment and clinical characteristics

This study was approved by the Ethics Committee of the University of Lübeck, and all procedures were conducted in accordance with the revised version of the Declaration of Helsinki. Written informed consent was obtained from all participants before study enrollment.

Participants were recruited from our outpatient clinics and from a pre-existing local cohort. Participants were classified into mutation carriers of the *HTT* gene expansion and HC. Given the challenge of differentiating between premanifest (pHD) and manifesting (mHD) patients with HD, mutation carriers were categorized based on the presence of overt motor symptoms.

All participants underwent assessment of general medical history, demographic data, genetic testing to characterize *HTT* CAG repeat length, and screening for magnetic resonance imaging (MRI) contraindications, previous illnesses, and the current medication status. The participants with less than 26 CAG repeats in *HTT* and an unremarkable neurological examination were considered mutation-free HC. The neurological examinations were videotaped and performed by trained movement disorders specialists. Neurological assessments were conducted in accordance with the Unified Huntington’s Disease Rating Scale (UHDRS), motor subscale, while cognitive function was evaluated using the Montreal Cognitive Assessment (MoCA). Depressive symptoms were assessed using the Beck Depression Inventory-II (BDI-II), which participants completed independently. Following clinical characterization, we performed a multimodal neuroimaging session, including T_1_-weighted structural imaging and ^31^P-MRSI of the basal ganglia and the cerebellum.

### Neuroimaging acquisition

All neuroimaging data were collected using a 3T Siemens MAGNETOM Skyra MRI scanner at the University of Lübeck, Center for Brain, Behaviour and Metabolism Core Facility. To exclude concomitant neurological conditions that were not previously diagnosed, all acquired scans were reviewed by experienced neuroradiologists. Only complete datasets were included in the following analyses.

### T_1_-weighted neuroimaging

Structural imaging was performed using a three-dimensional T_1_-weighted fast low-angle shot (fl3d) sequence with a 64-channel head-neck coil. The imaging protocol was optimized for volumetric analyses, employing the following parameters: voxel size = 1 × 1 × 1 mm^3^, field of view = 192 × 256 × 256 mm^3^, repetition time (TR) = 1900 ms, echo time (TE) = 2.44 ms, inversion time (TI) = 900 ms, flip angle = 9°, and a generalized autocalibrating partially parallel acquisition (GRAPPA) acceleration factor of two along the anterior/posterior phase encoding direction. The total scan time for the T_1_-weighted sequence was 5 minutes and 39 seconds.

### ^31^Phosphorus magnetic resonance spectroscopy imaging

^31^P-MRSI data were acquired using a double-tuned quadrature head coil (^1^H/^31^P, RAPID Biomedical) and a three-dimensional Chemical Shift Imaging (CSI) Free Induction Decay sequence. The acquisition parameters were optimized to balance spectral resolution and scan duration, using a voxel size of 30 × 30 × 30 mm^3^, a field of view of 240 × 240 × 240 mm^3^, TR = 2000 ms, TE = 2.3 ms, flip angle = 50°, sixfold weighted averaging, spectral bandwidth = 2000 Hz, vector size = 1024, Hamming filter width = 100%, and nuclear Overhauser effect disabled. Broadband proton decoupling was applied using the WALTZ-4 scheme. The total scan time for ^31^P-MRSI was 8 minutes and 4 seconds. To enhance the signal-to-noise ratio (SNR) within a clinically acceptable scan duration of less than 10 minutes, protocol parameters were optimized based on our previous studies.(17–19) The placement of the CSI grid and volumes of interest (VOI) was standardized, with two voxels positioned in the anterior basal ganglia (mainly covering the striatum) and two voxels placed in the cerebellum (Figure 1). The center of the VOI was positioned anterior to the anterior commissure, ensuring coverage of the caudate nucleus and anterior putamen while minimizing contamination from adjacent white matter. The borders of the VOI were adjusted to align parallel with the midline. The posterior boundaries of the voxel were carefully aligned with the anterior limb of the internal capsule to maintain the neuroanatomical focus on the striatum. For cerebellar voxel placement, the midline vermis and the fourth ventricle were used as anatomical references. The VOI was positioned bilaterally within the cerebellar hemispheres, ensuring inclusion of both gray and white matter regions while avoiding the tentorium cerebelli. The superior boundary was aligned with the primary fissure of the cerebellum, ensuring consistency in coverage across participants. Shimming adjustments were performed manually. A slightly larger shim volume than the predefined VOIs was selected.

**Figure 1.**
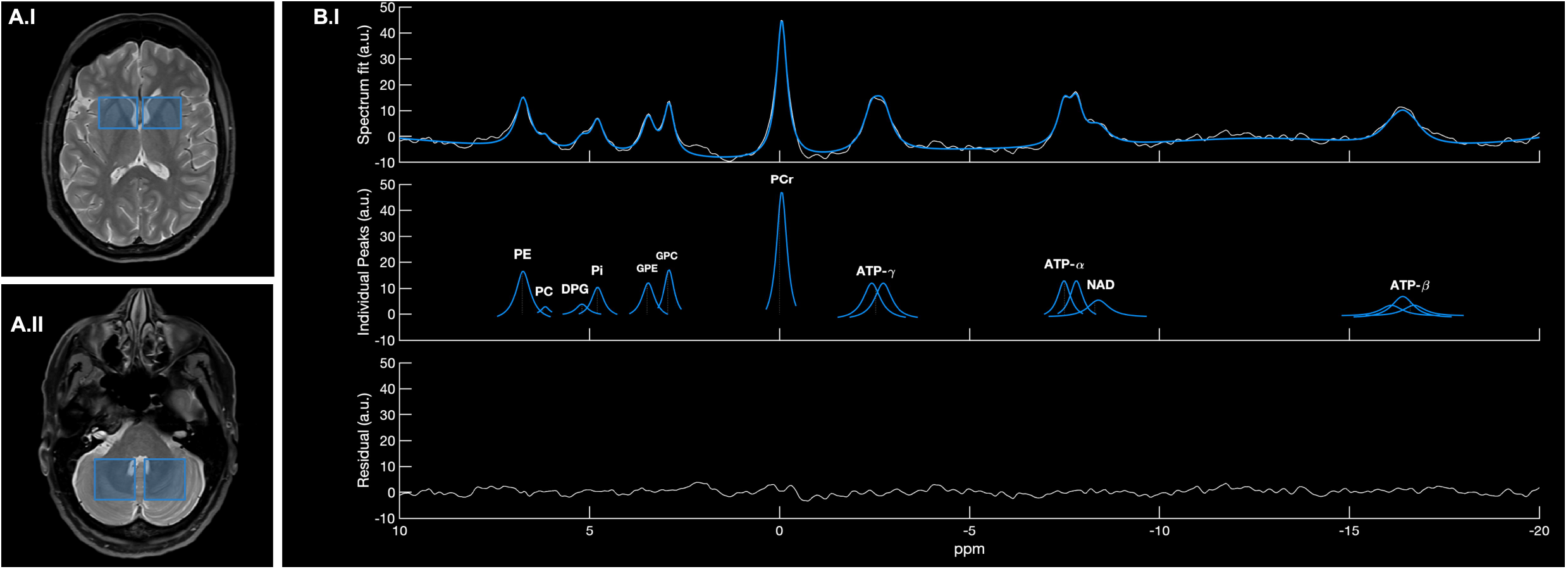
Region-specific voxel placement and representative ^31^phosphorus magnetic resonance spectroscopy imaging spectrum with postprocessing output. In panels A.I and A.II, we demonstrate anatomical localization of the acquisition voxel in the anterior basal ganglia (A.I) and cerebellum (A.II) on axial sections of the T_2_-weighted magnetic resonance images. The top section of panel B.I demonstrates the measured spectrum in white) overlaid with the fitted model (in blue). The middle section displays the individual metabolite peaks, and the bottom section demonstrates the residuals following spectral fitting. ATP-α, -β, -γ = adenosine triphosphate-alpha, -beta, -gamma; a.u. = arbitrary units; DPG = diphosphoglycerate; GPC = glycerophosphocholine; GPE = glycerophosphoethanolamine; NAD = nicotinamide adenine dinucleotide; PC = phosphocholine; PCr = phosphocreatine; PE = phosphoethanolamine; Pi = inorganic phosphate; ppm = particles per million.

### Neuroimaging postprocessing and analyses

Spectral analysis was performed using a Siemens workstation with the SynGo MR ER11 (VH22B_Sl19P26_CSI03) software package, following the procedures established in our previous reports.(17,19) This analysis pipeline included Fourier transformation with spatial zero-point filling, application of nuclear magnetic resonance prior knowledge, voxel-wise frequency shift correction, automatic phase correction, nonlinear curve fitting, and baseline correction. These steps ensured optimal spectral quality and accuracy in metabolite quantification. The quantification of HEPs was conducted by calculating metabolite ratios normalized to Pi. We defined (ATP-α+PCr)/Pi as the primary outcome variable. Additionally, we calculated ATP-α/Pi and PCr/Pi as the secondary outcome variables. To investigate whether the normalization procedure introduces any bias, we also report the above-stated metabolites without any referencing to Pi in an exploratory manner. Parameters derived from both neuroanatomical regions are reported as side-averaged mean values.

To account for potential confounding factors related to brain structure, volumetric data derived from T_1_-weighted imaging were included as potential covariates or correlating parameters. Volumetric analysis was conducted using AssemblyNet, a fully automated processing pipeline.(20) T_1_-weighted images underwent denoising, inhomogeneity correction, affine registration to MNI space, intensity normalization, and brain extraction to ensure standardized inputs. The segmentation framework employs a hierarchical ensemble of 3D U-Nets organized into two assemblies. The first assembly performs coarse segmentation at 2 × 2 × 2 mm^3^ resolution, which is refined by the second assembly at 1 × 1 × 1 mm^3^. Nearest neighbor transfer learning optimizes model initialization, and a majority voting mechanism aggregates the final segmentation, ensuring accuracy and robustness across datasets.(20) Total intracranial volume (TIV) was included as a covariate in the two-way analysis of covariance (ANCOVA) models, while the volumes of the caudate, globus pallidus, putamen, cerebellar white matter, cerebellar gray matter, and total cerebellar volume were considered as potentially correlating parameters in the analysis.

### Statistics

We conducted ANCOVA analyses to investigate group differences based on our primary and secondary outcome variables, accounting for age, sex, and TIV as covariates. Group (assignment to mHD, pHD, and HC) has been used as the fixed factor, and (ATP-α+PCr)/Pi, ATP-α/Pi, PCr/Pi, and ATP-α/PCr as the dependent variables analyzed separately for the anterior basal ganglia and the cerebellum. Post-hoc-comparisons were performed using Tukey’s range test. The same analysis was applied to the non-normalized metabolite levels and volumetric analyses. For volumetric analyses, we used TIV-corrected values for group comparisons (therefore, only accounted for age and sex as covariates) and non-TIV-corrected values as correlation parameters for correlative analyses with ^31^P-MRSI-derived metabolite data. Additionally, Spearman correlation analyses were conducted as an exploratory measure, with results reported uncorrected. All statistical analyses were performed in Jamovi (v.2.2.5.0) on a Microsoft Surface Pro 7 (Windows 11 Home), ensuring that all required assumptions were met before proceeding. Descriptive statistics are presented as mean ± standard deviation unless otherwise noted and are also expressed as ratios relative to the mean of the HC group to improve the interpretability of MRI-derived data. Graphical representations were generated using GraphPad Prism (v.10).

## RESULTS

### Demographics and clinical characteristics

The study included 15 mHD, 16 pHD, and 19 HC (Table 1). The distribution of sex was similar across groups: mHD: 8 females and 7 males, pHD: 10 females and 6 males pHD, and HC: 10 females and 9 males. The mHD were the oldest (58.41±18.3 years), followed by HC (50.8⍰ ±⍰17.6 years) and pHD (38.9⍰±⍰12.3 years), indicating reasonable age differences between groups. UHDRS motor score was markedly elevated in mHD (38.8⍰ ±⍰19.7), while pHD exhibited only subtle motor signs (2.1⍰±⍰2.1). *HTT* CAG repeat length was slightly higher in pHD (42.3⍰±⍰2.4) compared to mHD (41.5⍰±⍰1.6). For mHD, the average age of symptom manifestation was 51.6⍰±⍰8.4 years, and the mean disease duration was 7.9⍰±⍰5.4 years. MoCA scores were lower in mHD (20.5⍰±⍰5.3) compared to both pHD (26.6⍰±⍰2.6) and HC (26.6⍰±⍰2.3), with a statistically significant difference between the mHD and HC (p⍰< ⍰0.05). BDI-II scores were highest in mHD (13.9⍰±⍰8.6), followed by the intermediate in pHD (5.1⍰±⍰5.9), and lowest in HC (2.3⍰±⍰2.1). The difference in BDI-II scores between mHD and HC was statistically significant (p⍰< ⍰0.01).

**Table 1.**
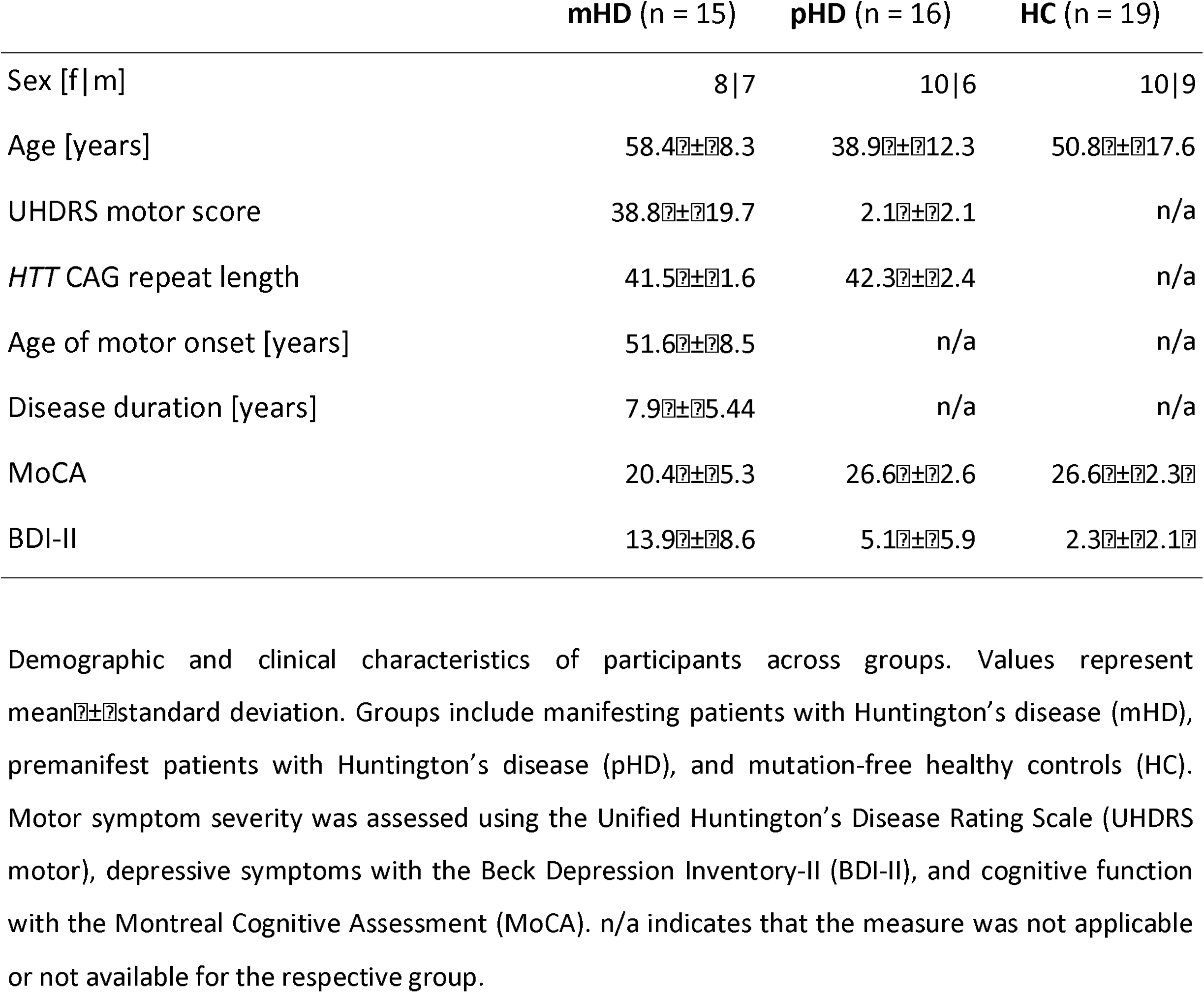
Demographic and clinical characteristics of study participants.

### Increased high-energy containing phosphates in the anterior basal ganglia of premanifest and manifest patients with Huntington’s disease

^31^P-MRSI revealed significant alterations in HEPs levels in the basal ganglia of *HTT* mutation carriers (Figure 2A and Table 2). The (ATP-α+PCr)/Pi ratio (F(2,44) ⍰= ⍰3.428, p⍰= ⍰0.041) was elevated in both mHD (8.09⍰±⍰1.85, +16.4%) and pHD (7.64⍰±⍰1.23, +10.0%) relative to HC (6.95⍰±⍰1.17), with a significant post hoc difference between mHD and HC only (p⍰= ⍰0.046). Non-normalized ATP-α+PCr (mHD: 1069.8⍰±⍰176.4, pHD: 1210.9⍰±⍰163.0, HC: 1086.6⍰±⍰203.0) and ATP-α (mHD: 496.6⍰±⍰93.3, pHD: 571.3⍰±⍰89.0, HC: 503.2⍰±⍰120.2) did not differ significantly between groups (p⍰> ⍰0.600). ATP-α/Pi ratio differed significantly across groups (F(2,44) ⍰=⍰3.415, p⍰=⍰0.042), mHD (3.78⍰±⍰1.10, +19.2%) and pHD (3.61⍰±⍰0.68, +13.9%) had higher values compared to HC (3.17⍰±⍰0.51), with a significant difference between mHD and HC only (p⍰= ⍰0.040). PCr/Pi showed no significant group differences (mHD: 4.30±0.80, pHD: 4.08⍰±0.85, HC: 3.78⍰±0.77, p = 0.069). Pi and PCr levels were comparable across all three groups (Pi: mHD: 137.2⍰±⍰33.5, pHD: 161.3⍰±⍰28.4, HC: 161.6⍰±⍰43.5, p⍰= ⍰0.094; and PCr: mHD: 573.2±95.5, pHD: 639.6±80.7, HC: 583.4±87.2, p = 0.572).

**Table 2.**
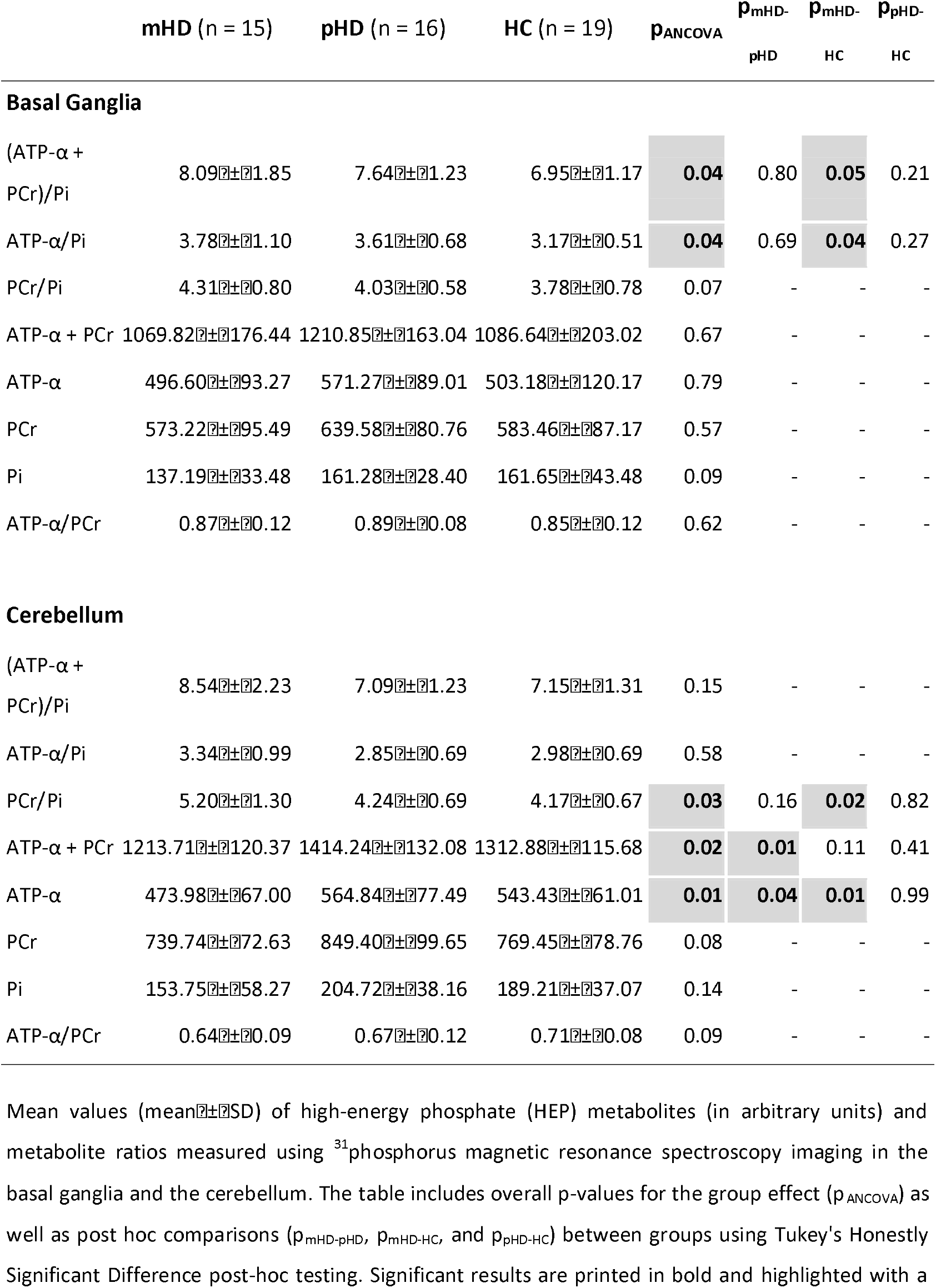
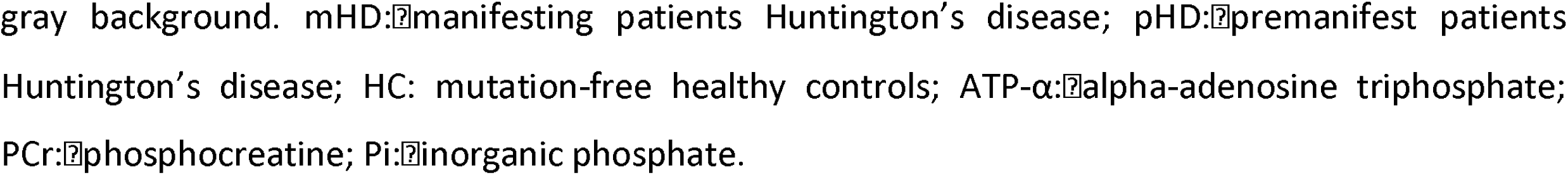
High-energy phosphate metabolites in the basal ganglia and cerebellum of patients with manifest, premanifest Huntington’s disease, and mutation-free healthy controls.

|  | mHD (n = 15) | pHD (n = 16) | HC (n = 19) | p<sub>ANCOVA</sub> | p<sub>mHD-pHD</sub> | p<sub>mHD-HC</sub> | p<sub>pHD-HC</sub> |
| --- | --- | --- | --- | --- | --- | --- | --- |
| **Basal Ganglia** |  |  |  |  |  |  |  |
| (ATP-α + PCr)/Pi | 8.09±1.85 | 7.64±1.23 | 6.95±1.17 | **0.04** | 0.80 | **0.05** | 0.21 |
| ATP-α/Pi | 3.78±1.10 | 3.61±0.68 | 3.17±0.51 | **0.04** | 0.69 | **0.04** | 0.27 |
| PCr/Pi | 4.31±0.80 | 4.03±0.58 | 3.78±0.78 | 0.07 | - | - | - |
| ATP-α + PCr | 1069.82±176.44 | 1210.85±163.04 | 1086.64±203.02 | 0.67 | - | - | - |
| ATP-α | 496.60±93.27 | 571.27±89.01 | 503.18±120.17 | 0.79 | - | - | - |
| PCr | 573.22±95.49 | 639.58±80.76 | 583.46±87.17 | 0.57 | - | - | - |
| Pi | 137.19±33.48 | 161.28±28.40 | 161.65±43.48 | 0.09 | - | - | - |
| ATP-α/PCr | 0.87±0.12 | 0.89±0.08 | 0.85±0.12 | 0.62 | - | - | - |
| **Cerebellum** |  |  |  |  |  |  |  |
| (ATP-α + PCr)/Pi | 8.54±2.23 | 7.09±1.23 | 7.15±1.31 | 0.15 | - | - | - |
| ATP-α/Pi | 3.34±0.99 | 2.85±0.69 | 2.98±0.69 | 0.58 | - | - | - |
| PCr/Pi | 5.20±1.30 | 4.24±0.69 | 4.17±0.67 | **0.03** | 0.16 | **0.02** | 0.82 |
| ATP-α + PCr | 1213.71±120.37 | 1414.24±132.08 | 1312.88±115.68 | **0.02** | **0.01** | 0.11 | 0.41 |
| ATP-α | 473.98±67.00 | 564.84±77.49 | 543.43±61.01 | **0.01** | **0.04** | **0.01** | 0.99 |
| PCr | 739.74±72.63 | 849.40±99.65 | 769.45±78.76 | 0.08 | - | - | - |
| Pi | 153.75±58.27 | 204.72±38.16 | 189.21±37.07 | 0.14 | - | - | - |
| ATP-α/PCr | 0.64±0.09 | 0.67±0.12 | 0.71±0.08 | 0.09 | - | - | - |

Mean values (mean ± SD) of high-energy phosphate (HEP) metabolites (in arbitrary units) and metabolite ratios measured using <sup>31</sup>phosphorus magnetic resonance spectroscopy imaging in the basal ganglia and the cerebellum. The table includes overall p-values for the group effect (p<sub>ANCOVA</sub>) as well as post hoc comparisons (p<sub>mHD-pHD</sub>, p<sub>mHD-HC</sub>, and p<sub>pHD-HC</sub>) between groups using Tukey's Honestly Significant Difference post-hoc testing. Significant results are printed in bold and highlighted with a
gray background. mHD: manifesting patients Huntington's disease; pHD: premanifest patients Huntington's disease; HC: mutation-free healthy controls; ATP-α : $\alpha$ -adenosine triphosphate; PCr: phosphocreatine; Pi: inorganic phosphate.

**Table 3.**
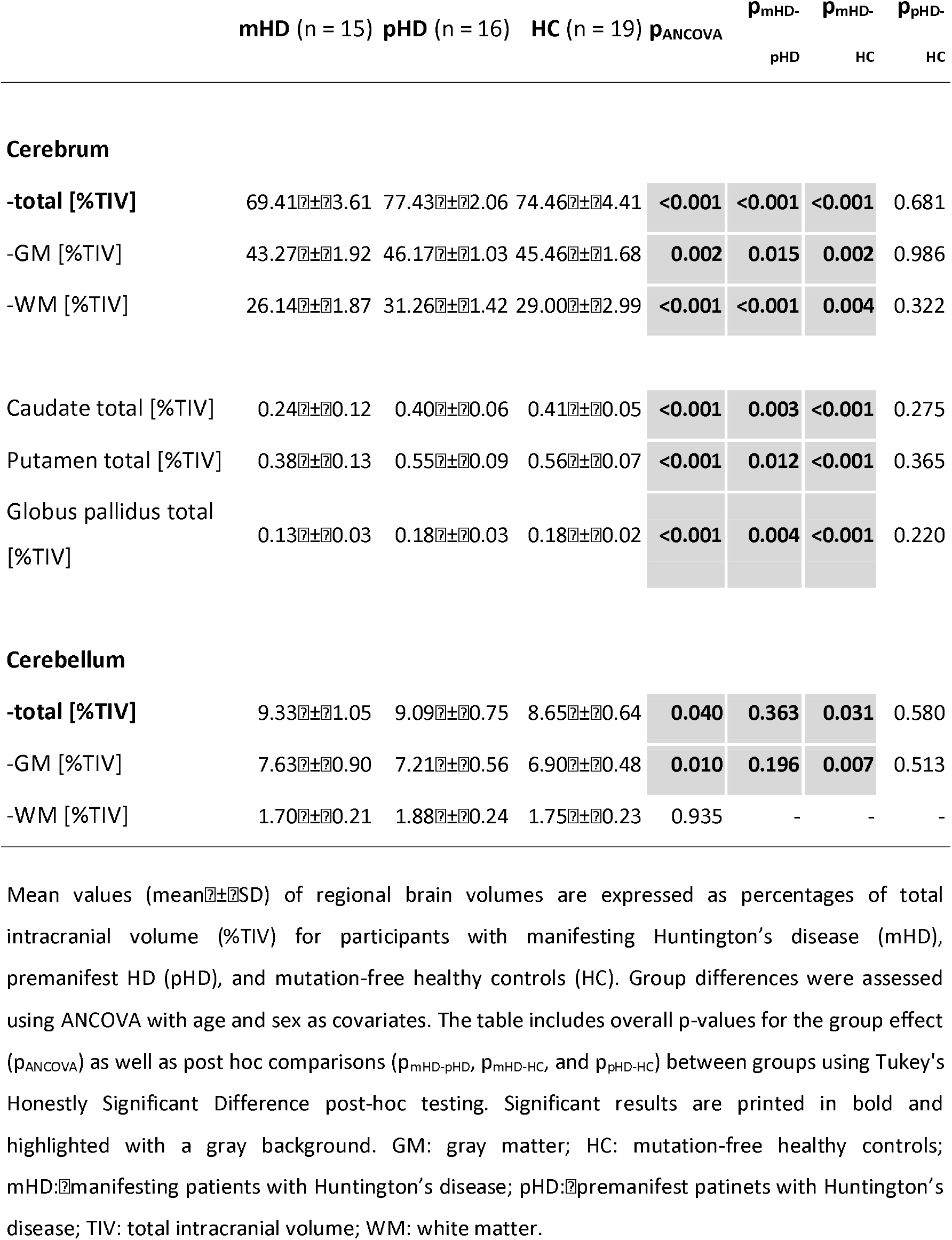
Regional brain volume differences in Huntington’s disease across disease-stages, cerebral, subcortical, and cerebellar structures.

**Figure 2.**
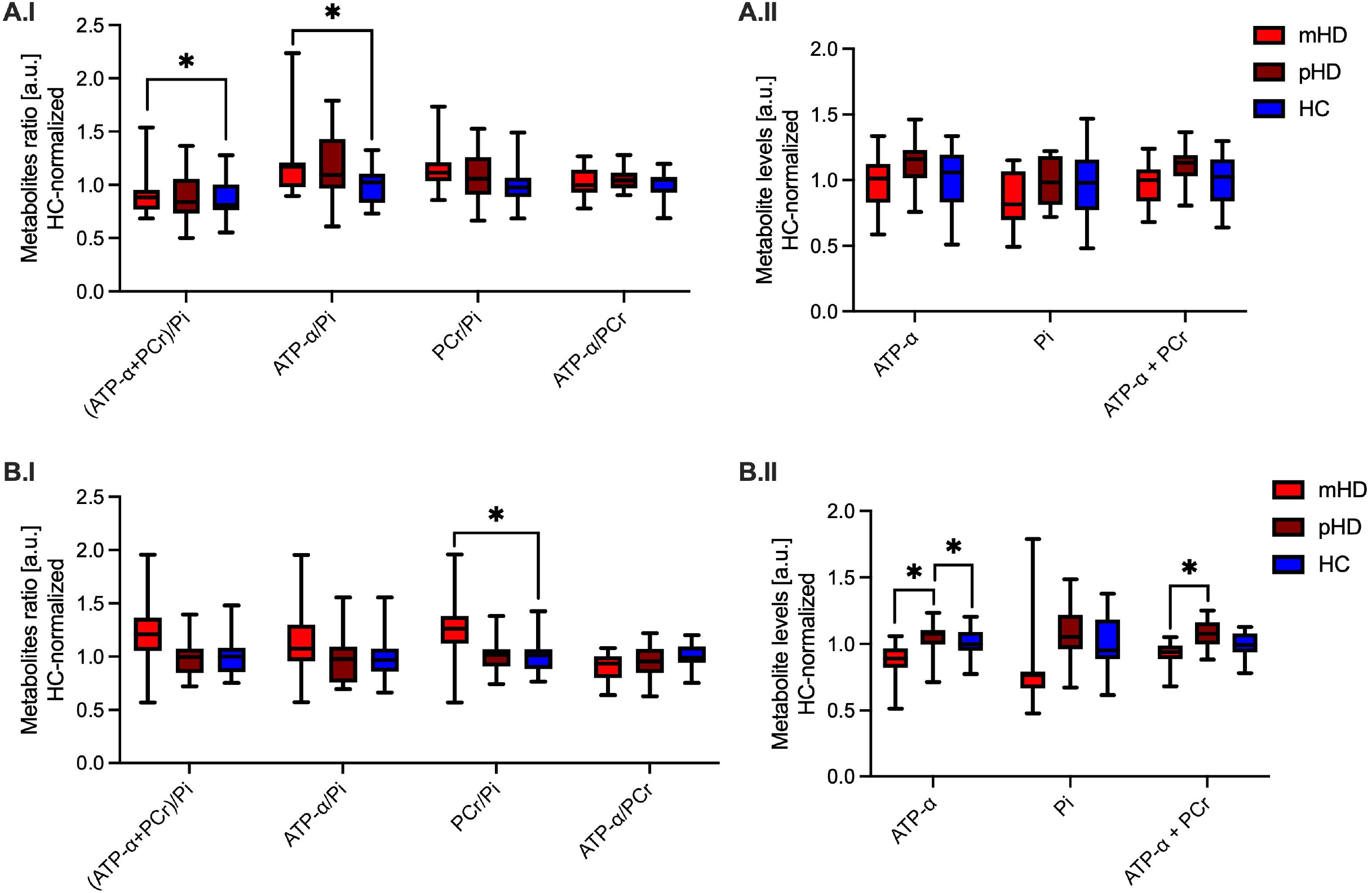
Region-specific box plot diagrams illustrating significant group differences in high-energy phosphate ratios across manifest, premanifest Huntington’s disease, and mutation-free healthy controls. Demonstrated values are derived from the two voxels in the anterior basal ganglia (panels A) and two voxels in the cerebellum (panels B), expressed in arbitrary units and normalized to the healthy control mean. In Panels A.I and B.I, we present metabolite ratios, and in panels A.II and B.II, the absolute levels of individual metabolites. Statistically significant group differences are indicated by brackets (* < 0.05). ATP-α = adenosine triphosphate phosphate alpha; a.u. = arbitrary units; HC = healthy controls; mHD = manifesting patients with Huntington’s disease; PCr = phosphocreatine; Pi = inorganic phosphate; pHD = premanifest patients with Huntington’s disease.

### Altered high-energy phosphate metabolism in the cerebellum of premanifest and manifest patients with Huntington’s disease

^31^P-MRSI revealed significant alterations in HEP levels in the cerebellum of HTT mutation carriers (Figure 2B). Although the (ATP-α+PCr)/Pi ratio showed a trend (mHD: 8.54⍰±⍰2.23, +19.5%; pHD: 7.09⍰±⍰1.23, −0.8%; HC: 7.15⍰±⍰1.31), the overall group effect was not statistically significant (F(2,44) ⍰= ⍰1.979, p⍰= ⍰0.150). The ATP-α/Pi ratio showed no significant differences across groups (F(2,44) ⍰= ⍰0.555, p⍰= ⍰0.578). We observed a significant group effect for the PCr/Pi ratio (F(2,44) ⍰= ⍰3.831, p⍰=10.029), with elevated levels in the mHD (5.20⍰±⍰1.30, +24.6%) and pHD (4.24⍰±⍰0.69, +1.7%) compared to HC (4.17⍰±⍰0.67). Post hoc comparisons confirmed a significant difference between mHD and HC (p⍰= ⍰0.024), while differences between pHD and HC were not significant (p⍰= ⍰0.824). ATP-α/PCr ratios across the three groups showed a trend (F(2,44) ⍰= ⍰2.610, p⍰= ⍰0.085), with no significant difference. Contrarily, ATP-α+PCr levels differed significantly across groups (F(2,44) ⍰= ⍰4.509, p⍰*=*10.017), with reduced values in mHD (1213.7⍰±⍰120.4, −7.6%) and elevated values in pHD (1414.2⍰±⍰132.1, +7.7%) relative to HC (1312.9⍰±⍰115.7). The pHD vs. mHD difference reached significance (p⍰= ⍰0.013). ATP-α levels demonstrated a significant group effect (F(2,44) ⍰= ⍰5.057, p⍰= ⍰0.011), with lower values in mHD (474.0⍰±⍰67.0, −12.8%) and higher values in pHD (564.8⍰±⍰77.5, +3.9%) compared to HC (543.4⍰±⍰61.0). Post hoc tests confirmed significant differences between mHD and pHD (p⍰= ⍰0.039) and between mHD and HC (p⍰= ⍰0.012). Pi levels (mHD: 153.8⍰±⍰58.3, pHD: 204.7⍰±⍰38.2, HC: 189.2⍰±⍰37.1; p⍰= ⍰0.138) did not differ significantly between groups.

### Stage-dependent volumetric changes in Huntington’s disease

Volumetric analysis revealed significant group differences in cerebral, subcortical, and cerebellar brain structures, with volumes expressed as a percentage of TIV, relative to the HC mean (Figure 3). Gray matter volume was reduced in mHD (43.27⍰±⍰1.92%) compared to HC (45.46⍰±⍰1.68%, p⍰= ⍰0.002; −4.8%) and pHD (46.17⍰±⍰1.03%, p⍰= ⍰0.015; −6.3%). Total cerebral volume in mHD was 69.41⍰±⍰3.61%, showing a −6.8% reduction versus HC (74.46⍰±⍰4.41%, p⍰< ⍰0.001) and a −10.4% reduction relative to pHD (77.43⍰±⍰2.06%, p⍰< ⍰0.001). Cerebral white matter volume was significantly lower in mHD (26.14⍰±⍰1.87%) compared to HC (29.00⍰±⍰2.99%, −9.9%, p1=10.004) and pHD (31.26⍰±⍰1.42%, −16.4% p⍰< ⍰0.001). No significant cerebral volume differences were observed between pHD and HC (ps⍰> ⍰0.300). Caudate volume was reduced by −40.0% in mHD (0.243⍰±⍰0.116%) compared to HC (0.405⍰±⍰0.046%, p⍰< ⍰0.001), and by −39.9% compared to pHD (0.3951±10.063%, p1=10.003). Globus pallidus volume was −30.8% lower in mHD (0.127⍰±⍰0.031%) than in HC (0.184⍰±⍰0.021%, p⍰< ⍰0.001), and −29.4% lower than in pHD (0.180⍰±⍰0.031%, p⍰= ⍰0.004). Similarly, putamen volume in mHD (0.378⍰±⍰0.134%) was reduced by −32.4% versus HC (0.559⍰±⍰0.065%, p⍰< ⍰0.001) and −31.7% versus pHD (0.554⍰±⍰0.093%, p⍰= ⍰0.012). No significant differences were found between pHD and HC for any basal ganglia structure (ps⍰> ⍰0.200).

**Figure 3.**
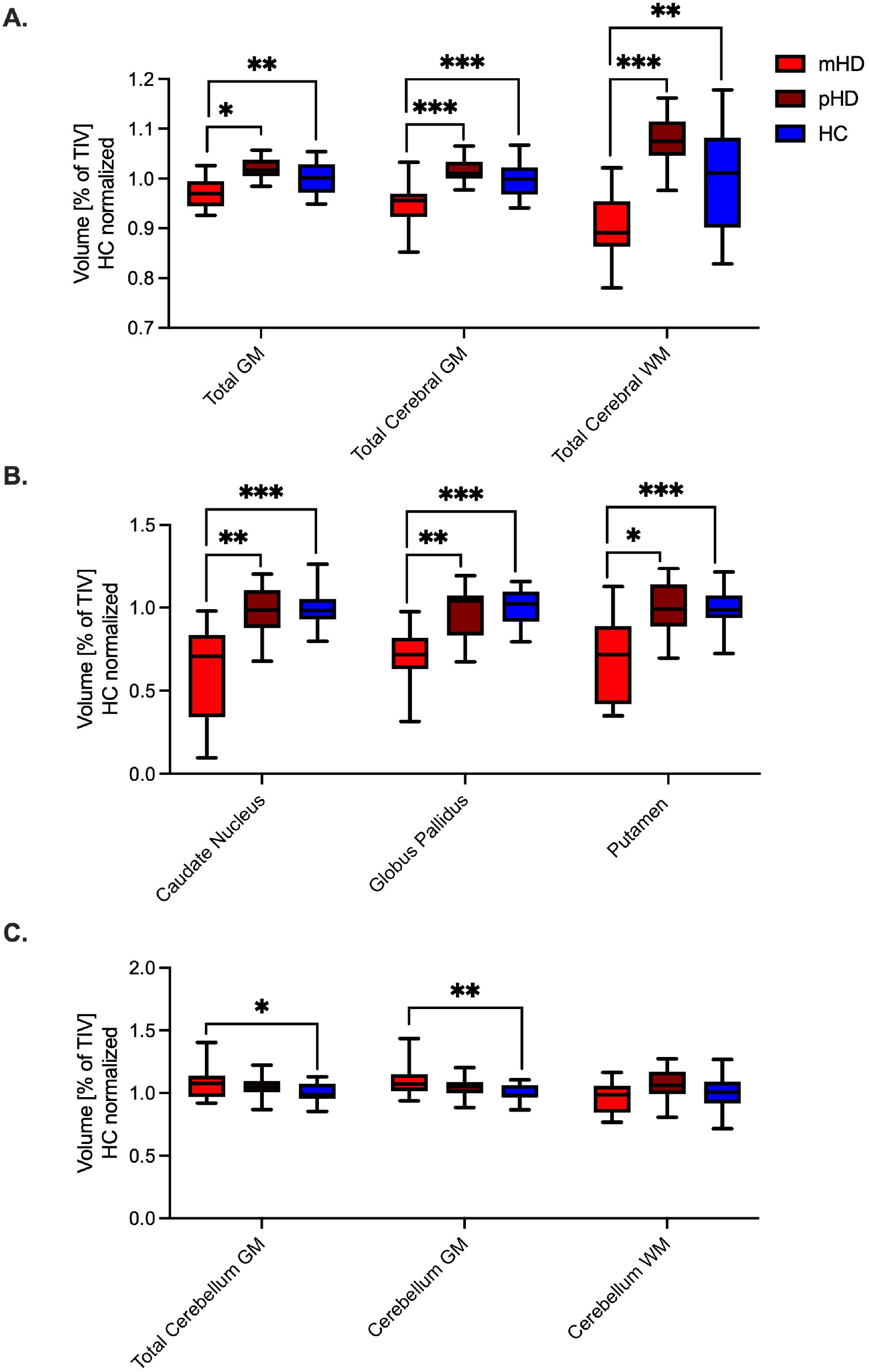
Group comparisons of cerebral, subcortical, and cerebellar volumetric measures across premanifest and manifesting patients with Huntington’s disease compared with mutation-free healthy controls. Brain volumes are presented as a percentage of total intracranial volume and normalized to the healthy control mean. The cerebral (panel A), subcortical (B), and cerebellum (C) volumetric measures from premanifest (dark red) and manifesting (light red) patients with Huntington’s disease and healthy controls (blue) are presented. The significant group differences are indicated by brackets (*** < 0.001; ** < 0.01; * < 0.05). GM = gray matter; HC = healthy controls; mHD = manifesting patients with Huntington’s disease; pHD = premanifest patients with Huntington’s disease; TIV = total intracranial volume; WM = white matter.

**Figure 4.**
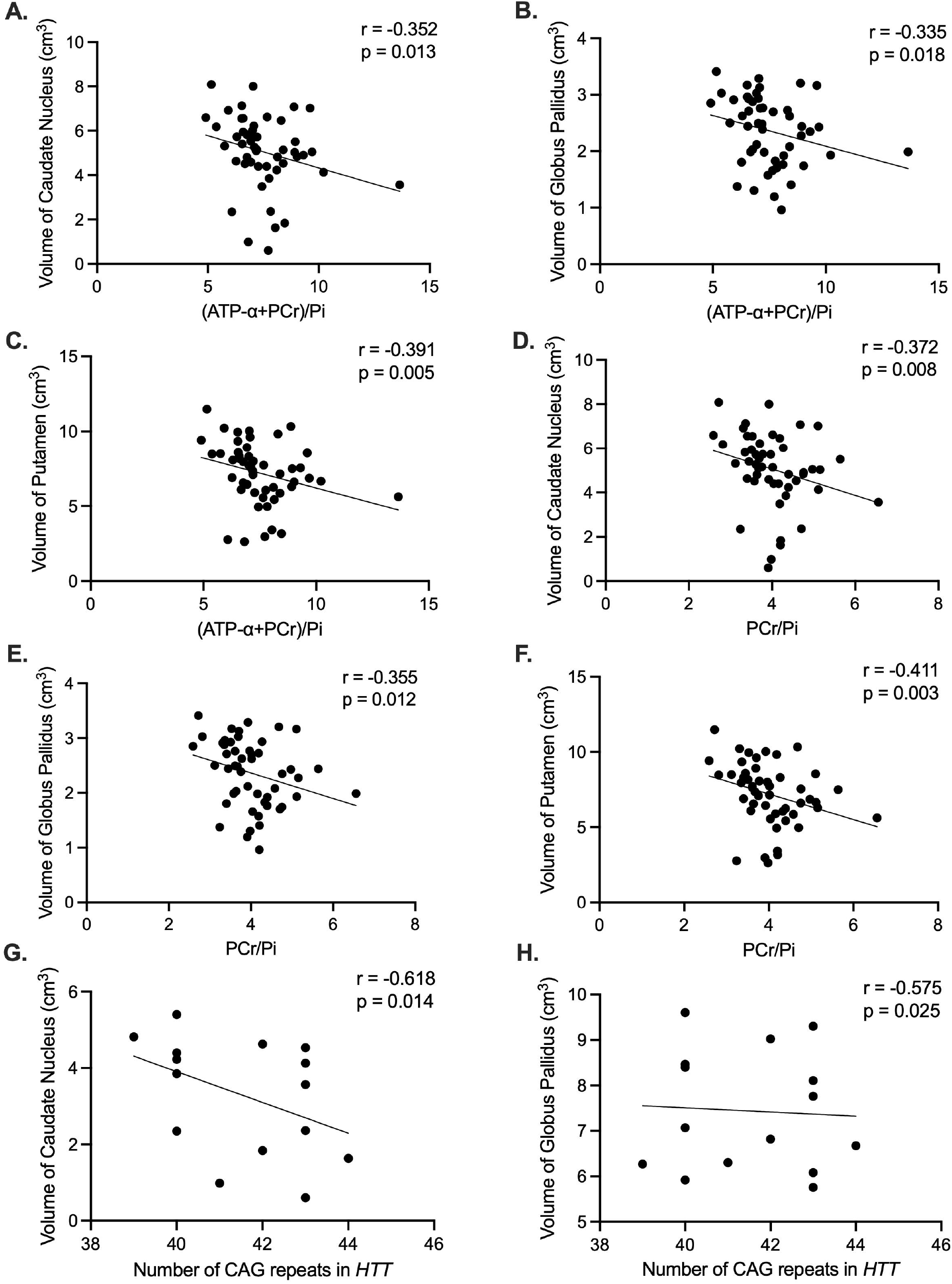
Associations between basal ganglia volumes and high-energy phosphate metabolites ratios and the number of CAG repeats in the Huntington gene. We present scatter plots demonstrating the relationship between regional basal ganglia volumes and (ATP-⍰+PCr)/Pi (panels A-C), PCr/Pi (panels D-F), and the number of CAG repeats in the mutant Huntington gene. ATP-1 = adenosine triphosphate alpha; *HTT* = mutant Huntington gene; p = p-value; PCr = phosphocreatine; Pi = inorganic phosphate; r = correlation coefficient.

Total cerebellar volume was modestly increased in mHD (9.333⍰±⍰1.052%, +7.9%) relative to HC (8.653⍰±⍰0.637%), with a significant difference between mHD and HC (p⍰= ⍰0.031), while pHD individuals showed a +5.0% increase (9.087⍰±⍰0.750%) and no significant with HC (p⍰= ⍰0.580) or mHD (p⍰= ⍰0.363). Cerebellar gray matter volume demonstrated a significant group effect (F(2,45) ⍰=15.087, p⍰= ⍰0.010), with mHD participants showing a +10.6% increase (7.632⍰±⍰0.899%) compared to HC (6.899⍰±⍰0.480%, p⍰= ⍰0.007). The pHD group had intermediate values (7.210⍰±⍰0.564%), which did not differ significantly from mHD (p⍰= ⍰0.196) or HC (p⍰= ⍰0.513). In contrast, cerebellar white matter volume was not different across all groups (F(2,45) ⍰= ⍰0.067, p⍰= ⍰0.935), with values of 1.702⍰±⍰0.214% in mHD, 1.877⍰±⍰0.237% in pHD, and 1.754⍰±⍰0.234% in HC.

### Associations between high-energy-containing phosphates and brain volumes

Higher HEP ratios were consistently associated with smaller volumes of basal ganglia nuclei. Specifically, the (ATP-α+PCr)/Pi ratio negatively correlated with caudate (rho⍰= ⍰-0.352, p=⍰0.013), globus pallidus (rho =⍰-0.335, p =⍰0.018), and putamen volumes (rho⍰= ⍰-0.391, p⍰= ⍰0.005). Moreover, the ATP-α/Pi ratio demonstrated significant negative correlations with caudate (rho1=1-0.281, p⍰= ⍰0.049) and putamen volumes (rho⍰= ⍰-0.310, p⍰= ⍰0.029). Similarly, the PCr/Pi ratio was inversely associated with caudate (rho =⍰-0.372, p⍰= ⍰0.008), globus pallidus (rho⍰= ⍰-0.355, p⍰= ⍰0.012), and putamen volumes (rho =⍰-0.411, p⍰= ⍰0.003). Contrarily, cerebellar correlations were less consistent and weaker. While the PCr/Pi ratio in the cerebellum was negatively associated with cerebellar white matter volume (rho⍰= ⍰-0.320, p⍰= ⍰0.024), other metabolite ratios, including (ATP-α+PCr)/Pi, ATP-α/Pi, and ATP-α/PCr, did not demonstrate significant associations with cerebellar gray or total volume.

### Association between high-energy-containing phosphates and volumetric measures with clinical scores and genetic parameters

Across all regions of interest and in both pHD and mHD, (ATP-α+PCr)/Pi, ATP-α/Pi, PCr/Pi, or ATP-α/PCr ratios showed no significant associations with UHDRS motor scores, disease duration, *HTT* repeat length, BDI-II, or MoCA scores (all p⍰> ⍰0.100). *HTT* CAG repeat length was negatively correlated with caudate volume (rho⍰= ⍰-0.618, p1=10.014) and globus pallidus volume (rho⍰= ⍰-0.575, p⍰=10.025) in mHD. A similar trend was observed for the putamen (rho⍰= ⍰-0.428), although this did not reach statistical significance (p⍰= ⍰0.111). Contrarily, no significant correlations between repeat length and subcortical volumes were observed in pHD (all p⍰> ⍰0.600). When analyzing all *HTT* mutation carriers together (pHD⍰+ ⍰mHD), repeat length was not significantly associated with striatal, cerebellar, or cerebral volumes (all p⍰> ⍰0.300). No significant associations were found between *HTT* repeat length and cerebellar volumes (total, gray matter, or white matter) nor with cerebral gray or white matter volumes, in either group.

## DISCUSSION

This study provides novel evidence for region- and stage-specific alterations in cerebral and cerebellar HEP metabolism in *HTT* mutation carriers, using ^31^P-MRSI. We observed significantly elevated (ATP-α + PCr)/Pi and ATP-α/Pi ratios in the basal ganglia and PCr/Pi in the cerebellum of mHD compared to HC. While non-normalized metabolite concentrations showed a stage-dependent pattern in the cerebellum, being elevated in the pHD and reduced in the mHD. These metabolic changes were not associated with clinical scores but showed consistent negative correlations with the volume of striatal gray matter structures, suggesting a structural-metabolic coupling that may precede progression into the overt clinical disease.

The basal ganglia structures are one of the earliest and most severely affected regions in HD, with progressive neuronal loss, synaptic dysfunction, and mitochondrial impairment.(21) The increased HEP ratios in pHD may reflect compensatory upregulation, in which mitochondrial activity and creatine kinase buffering systems are transiently enhanced to maintain neuronal function despite early synaptic or oxidative stress. This is consistent with the observation of non-significantly altered non-normalized concentrations of ATP-α, PCr, and Pi, suggesting that energy buffering efficiency, rather than the total energy pool size, is altered in pHD. As the disease progresses to the mHD stage, sustained metabolic stress may lead to decompensation, resulting in a relative energy deficit, reflected by altered metabolite ratios and declining absolute concentrations, specifically in the cerebellum.

The stage-dependent observations in the cerebellum, with increased ATP-α and PCr levels in pHD and reduced levels in mHD, indicate that the cerebellum initially mounts a metabolic resilience response, possibly through astrocytic support or enhanced oxidative phosphorylation, before succumbing to broader network dysfunction and energetic collapse as the disease advances. The reduced ATP-α/PCr ratio observed across mutation carriers suggests a shift in the balance between ATP production and creatine buffering, possibly reflecting mitochondrial inefficiency or impaired substrate utilization. The observation of higher cerebellar gray matter volume in mHD may present a compensatory structural response to the observed metabolic dysregulation. However, previous research has reported heterogeneous findings regarding cerebellar volume in HD, including widespread atrophy and increased volumes.(22) Further, accumulating evidence demonstrates the role of the cerebellum in early stages of the disease, potentially occurring independently of striatal degeneration, contributing to the disease onset.(7)

Importantly, the observed metabolic alterations were not associated with clinical severity, suggesting that bioenergetic dysregulation is possibly an early event in HD pathogenesis, preceding clinical manifestations. Simultaneously, the inverse correlation between HEP ratios and striatal volumes highlights a structure-function coupling, whereby atrophic regions exhibit greater energy stress or compensatory metabolic activity, consistent with findings from other neurodegenerative diseases.(18)

Our study has several limitations, such as a cross-sectional design, which limits causal inference regarding the temporal evolution of HEP-related metabolism across disease stages; longitudinal studies are needed to determine whether the observed bioenergetic shifts predict clinical progression or structural decline. Although we employed state-of-the-art ^31^P-MRSI at 3T, spatial resolution remains limited, particularly in small subcortical structures such as the globus pallidus or in cerebellar lobules, which may have led to partial volume effects. While our sample included both pHD and mHD individuals, the relatively modest group sizes and limited range of pathologic *HTT* CAG repeats may limit statistical power, especially for detecting subtle correlations with clinical variables. Additionally, ^31^P-MRSI cannot directly resolve mitochondrial vs. other (e.g., glycolytic) ATP sources or distinguish neuronal from glial contributions to the measured signals.

Future research should aim to validate these findings in larger, longitudinal cohorts and integrate ^31^P-MRSI with complementary imaging modalities such as PET imaging or tracer-based ^13^C-MRSI to better characterize the cellular and molecular underpinnings of energy metabolism in HD. Moreover, coupling metabolic imaging with blood-based or cerebrospinal fluid biomarkers of mitochondrial function, oxidative stress, or neuroinflammation could help delineate causal pathways. Finally, ^31^P-MRSI may serve as a valuable translational readout for evaluating emerging metabolic therapies, including creatine, NAD+ precursors, or mitochondrial-targeted antioxidants, particularly in the pHD where therapeutic intervention is most promising.(1,13,23)

Taken together, our results suggest a dynamic, regionally heterogeneous pattern of disturbed HEP metabolism in HD, characterized by early compensatory changes in HEP buffering in vulnerable brain regions, followed by energetic decline specifically in the cerebellum with disease progression. This highlights the potential of ^31^P-MRSI not only as a tool to monitor early pathophysiological changes in HD but also as a platform for evaluating future metabolic interventions aimed at restoring bioenergetic homeostasis.

## ACKNOWLEDGMENT

We would like to thank our study nurse, Jenny Schmalfeld, for her commitment, meticulous organization, and continuous support throughout participant recruitment, study coordination, and data acquisition. We are also deeply grateful to all patients who participated in this study, as well as to their caregivers, for their time, trust, and invaluable contribution, without which this research would not have been possible.

## AUTHOR CONTRIBUTION

**JP:** Conceptualization, Methodology, Investigation, Formal analysis, Data curation, Visualization, Writing original draft, Supervision. **MGKM:** Conceptualization, Methodology, Investigation, Formal analysis, Visualization, Writing original draft. **MMP:** Investigation, Data curation, Validation, Visualization, Writing review and editing. **JLA:** Investigation, Data curation, Writing review and editing. **LvW:** Investigation, Data curation, Writing review and editing. **JH:** Investigation, Data curation, Writing review and editing. **JU:** Investigation, Data curation, Writing review and editing. **SL:** Investigation, Data curation, Writing review and editing. **AM:** Resources, Clinical expertise, Writing review and editing. **NB:** Conceptualization, Supervision, Project administration, Funding acquisition, Writing review and editing.

## DATA AVAILABILITY

The data that supports the findings of this study will be made available upon reasonable request from the corresponding author.

